# Antibody Response Following COVID-19 Boosters During the Omicron Wave in the United States: A Decentralized, Digital Health, Real-World Study

**DOI:** 10.1101/2022.07.31.22278173

**Authors:** Tosha Doornek, Nan Shao, Paul Burton, Francesca Ceddia, Belen Fraile

**Author notes:** **Corresponding Author Information:** Belen Fraile, Address: 200 Technology Square, Cambridge, MA 02139, USA.

## Abstract

As the coronavirus disease 2019 (COVID-19) pandemic evolves, new methods that enable real-world studies can provide public health information regarding long-term immunity following vaccination and booster campaigns. We conducted an observational trial to assess antibody levels against SARs-CoV-2 following COVID-19 vaccine boosters in adults in the United States during the omicron wave. This study utilized mobile health technology to recruit and enroll participants in combination with at-home blood collection self-conducted by participants and self-administration of surveys. The results show that when compared with the Pfizer-only primary/booster COVID-19 vaccine, the Moderna-only primary/booster vaccine substantially boosts and maintains high antibody titers over time against the ancestral strain and variants, including BA.1; the geometric mean ratios were generally statistically significantly greater when comparing the Moderna-only to the Pfizer-only vaccine/booster series over much of the study period. These observations suggest that priming/boosting with the Moderna vaccine may be highly protective against COVID-19; such data are vital to inform future recommendations for COVID-19 boosters and to assess the emergence of variants.

## Main Body

As the COVID-19 pandemic evolves, it is necessary to track and describe immune responses among COVID-19 vaccine recipients in real-world settings. These data can help health systems, policy makers, and regulators, make public health decisions around vaccination campaigns and assess the emergence of variants. The real-world characterization of antibody levels against SARS-CoV-2 over time following administration of different booster regimens is unknown. We employed an innovative methodology that used mobile health technology to recruit/enroll participants, combined with an at-home participant-facilitated blood collection device, to conduct a trial assessing SARS-CoV-2 antibody levels following COVID-19 boosters during the omicron wave (NCT05367908). To our knowledge, this study is the first of its kind to utilize decentralized procedures to assess real-world immunogenicity following COVID-19 vaccination.

Participants were identified/enrolled through the digital health community Evidation (Evidation Health, California). US adults ≥18 years were eligible for the study if they received a COVID-19 booster between September 2021 and screening, and their self-reported information was confirmed to have met all inclusion/exclusion criteria. Participant vaccination cohorts (confirmed by vaccination cards) were MMM (Moderna-only primary/booster) or PPP (Pfizer-only primary/booster). Overall, 844 participants were included (MMM, n=378; PPP, n=466), with a mean age of 44±13 years (**Table S1**). Participants received their first booster 1 to 7 months before study entry. Participants collected blood using the YourBio TAP II device (Medford, Massachusetts)^1^ at 0, 1, and 2 months post-enrollment and mailed samples for laboratory-based antibody measurement using the MSD VAC123 assay. Numerically higher geometric mean antibody titers against the ancestral strain and variants of concern (VOCs), including BA.1, were observed with MMM vs PPP from ≥8 through ≥32 weeks following first booster (**Figure 1**). MMM titers remained stable for 20 weeks following first booster, followed by a decline, whereas PPP titers decreased starting from 8 weeks following first booster, suggesting longer antibody retention among MMM recipients. Generally, geometric mean ratios were statistically significantly greater than 1 when comparing MMM against PPP from ≥12 weeks through ≥32 weeks following first booster.

**Figure 1.**
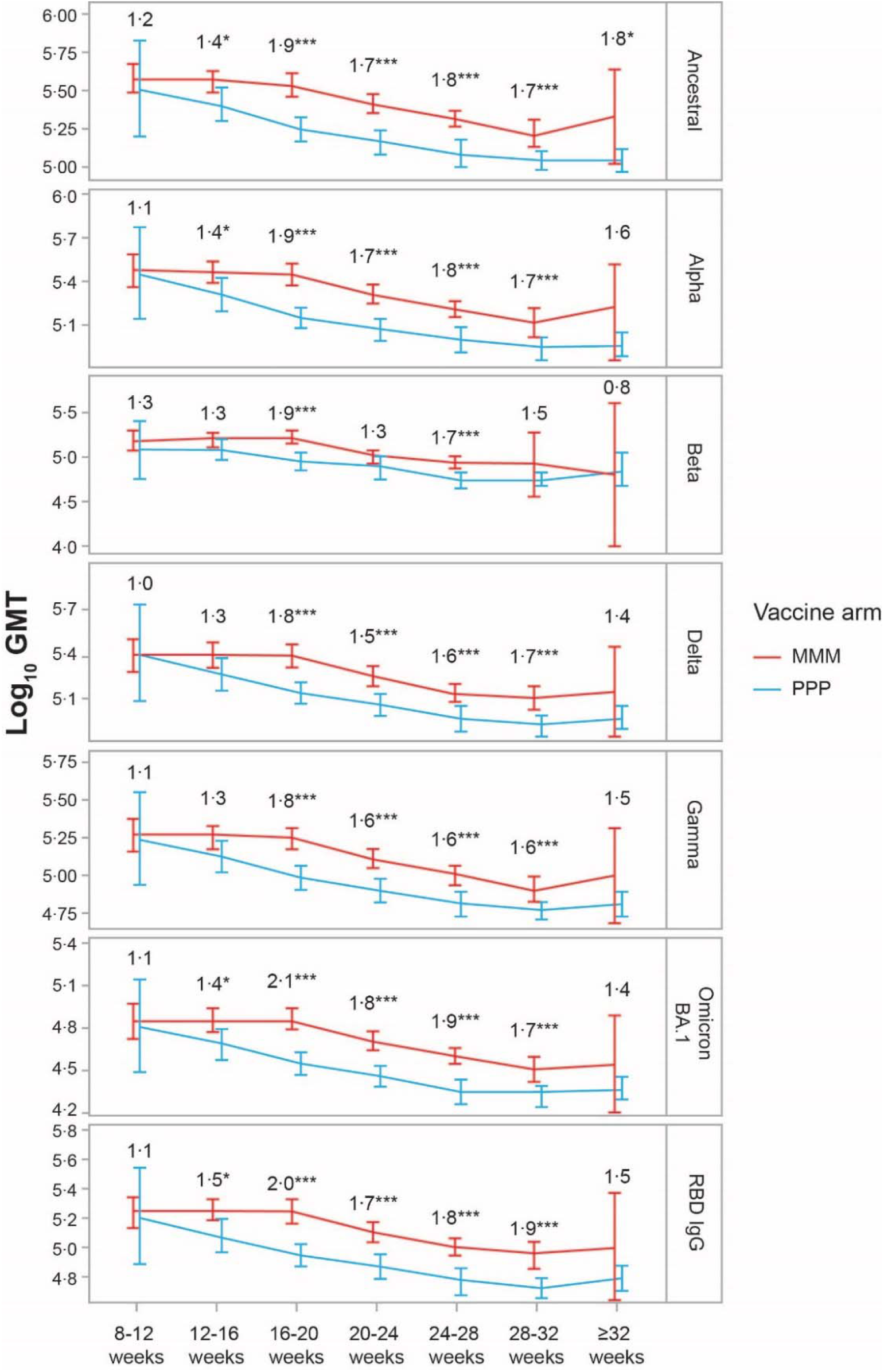
Antibody GMTs and GMRs From ≥8 Weeks Through ≥32 Weeks Following First Booster in the MMM and PPP Cohorts. GMTs and corresponding 95% CIs were based on raw data without model adjustment and were plotted over time for the MMM and PPP vaccine/booster series arms. The number displayed above each GMT represents the GMR when comparing MMM vs PPP, calculated from the ANCOVA model. The presence of an asterisk next to a value represents the statistical significance level of the GMR: ***, p-value ≤0□001; **, p-value >0□001 and ≤0□01; *, p-value >0□01 and ≤0□05; no symbol, p-value >0□05. Antibody results from 0 through 8 weeks following first booster are not displayed in the figure, as there were insufficient data (n=2 participants each in the MMM and PPP cohorts). ANCOVA, analysis of covariance; GMR, geometric mean ratio; GMT, geometric mean titer; IgG, immunoglobulin G; MMM, Moderna-only vaccine/booster series; PPP, Pfizer-only vaccine/booster series; RBD, receptor binding domain.

The innovative methodology employed here sets the precedent for greater research access using a digital platform and consumer-directed technology and serves as a model for real-world studies requiring timely data collection. The study demonstrated that the Moderna-only primary/booster COVID-19 vaccine substantially boosts and maintains high antibody titers over time against the ancestral strain and VOCs, including BA.1, suggesting that priming/boosting with this vaccine may be highly protective against COVID-19.

## Data Availability

All data produced in the present study are available upon reasonable request to the authors.

## Acknowledgements

The authors thank the participants for their participation in this study. Medical writing and editorial assistance were provided by Kate Russin, PhD, of MEDiSTRAVA in accordance with Good Publication Practice (GPP3) guidelines, funded by Moderna, Inc., and under the direction of the authors.

## Author Contributions

PB, TD, and NS were involved in the study design. Data collection was performed by TD. Data analysis and interpretation were undertaken by NS. BF, FC, PB, TD, and NS were involved in the drafting and critical review of the manuscript. All authors give final approval of the published version and agree to be accountable for all aspects of the work.

Tosha Doornek and Nan Shao verify that they had access to all the data and take responsibility for it.

