## Supplement for "Antibody Response Following COVID-19 Boosters During the Omicron Wave in the United States: A Decentralized, Digital Health, Real-World Study"

### Supplemental Methods

#### Study Objectives

The primary objective of the study was to describe changes in antibody levels among US adults over time following receipt of a COVID-19 booster, using an at-home capillary blood draw device to collect a serum sample for subsequent antibody testing in a decentralized manner. The secondary objectives of the study were to describe changes in antibody levels over time between subpopulations of interest (eg, age groups, vaccine type, COVID infection), where possible, and to describe changes in antibody levels to various COVID-19 variants over time since receiving a COVID-19 booster.

#### Ethics Statements

The protocol, amendments, informed consent form, recruitment materials, and all participant materials were submitted to the Advarra Institutional Review Board (Columbia, Maryland, United States) for review and approval. Approval of these documents was obtained before implementation of study procedures.

#### Participant Inclusion/Exclusion Criteria

Participants who met all of the following criteria were eligible for enrollment into the study:

- ≥18 years
- Live in the continental United States
- Can speak, read, and understand English
- Vaccinated against COVID-19 with a US-authorized vaccine (ie, received the single-shot Johnson & Johnson viral vector vaccine or both shots in the Pfizer or Moderna mRNA vaccine series; of note, this publication reports results for those who received Moderna-only primary/booster and Pfizer-only primary/booster)
- Received a US-authorized COVID-19 booster between September 2021 and screening
- Willing and able to submit vaccination card photo(s)
- Willing and able to self-collect capillary blood 3 times during the study period via an at-home whole-blood collection device (ie, the YourBio TAP II device)

Participants who meet any of the following criteria were excluded from the study:

- Diagnosed with significant cognitive impairment or dementia
- Received >1 COVID-19 vaccine booster at screening
- Pregnant or planning to become pregnant during the study period
- Primary mailing address is a PO box, Army Post Office (APO), Fleet Post Office (FPO), or Diplomatic Post Office (DPO) address
- Currently live in a US military base located overseas, or US territories (Puerto Rico, US Virgin Islands, Guam, Northern Mariana Island, or American Samoa)
- Currently participating in a COVID-19 vaccine clinical trial
- Currently receiving chemotherapy or has received chemotherapy in the past 6 months
- Currently taking steroids, such as prednisone, for any condition
- Diagnosed with or is taking medications for rheumatoid arthritis, lupus, or multiple sclerosis
- Received an organ transplant
- Currently undergoing dialysis of any kind (for example, hemodialysis or chronic ambulatory dialysis) for kidney disease

#### Statistical Analyses

No formal power analysis was conducted to determine the sample size, given the primary objective was exploratory in nature. The final data cut for this interim analysis was July 5, 2022. The primary analysis model to evaluate the geometric mean ratio and statistical comparisons between cohorts was an analysis of covariance (ANCOVA) model, with log-transformed antibody level as the dependent variable and vaccine type as the fixed effect, adjusting for age group (18-25; 26-64; ≥65 years) and COVID-19 infection prior to study entry (yes/no). Separate ANCOVA models were built for each time since first booster interval. At approximately 1, 2, and 3 months following study enrollment, participants were asked to self-report, via an online survey, the following criteria in relation to their last completed survey:

- Tested positive for COVID-19
- Received a second COVID-19 vaccine booster
- Started receiving chemotherapy
- Started taking steroids, such as prednisone, for any health condition
- Received a diagnosis of or started taking any new medications for rheumatoid arthritis, lupus, or multiple sclerosis
- Started dialysis of any kind (for example, hemodialysis or chronic ambulatory dialysis) for kidney disease

Participants who answered “yes” to the above questions were censored for the analysis (ie, corresponding antibody values were considered as missing). In cases where the date of the event was known, censoring was applied from the date of the event. In cases where the event date was unknown, censoring was applied to the date of the monthly survey immediately before the survey where the criterion was self-reported. Differences in GMRs between cohorts were considered statistically significant at p-values ≤0⸱05.

### Supplemental Results

#### Participants

Most participants were female (60%), and the mean body mass index was 28±6 kg/m^2^ in the overall sample population (**Table S1**). Participants were representative of a diverse population, with 48% reporting as a race/ethnicity other than White.

#### Limitations

Limitations of this study include the reduced sample size as a result of the recommendation by the CDC for an additional booster in combination with the COVID-19 surge. Additionally, as this was a novel study design, anticipating and preparing for issues that could develop during the study was challenging.

### Supplemental Tables

#### Table S1. Participant Demographics and Baseline Characteristics

|  | **MMM**  **(n=378)** | **PPP**  **(n=466)** | **Overall**  **(N=844)** |
| --- | --- | --- | --- |
| **Age group, years** | | | |
| ≥65 | 44 (11⸱6%) | 42 (9⸱0%) | 86 (10⸱2%) |
| 18-25 | 16 (4⸱2%) | 26 (5⸱6%) | 42 (5⸱0%) |
| 26-64 | 318 (84⸱1%) | 398 (85⸱4%) | 716 (84⸱8%) |
| **Age at enrollment, years** | | | |
| Mean (SD) | 44⸱8 (13⸱5) | 43⸱3 (13⸱1) | 44⸱0 (13⸱3) |
| Median [Min, Max] | 42⸱0 [19⸱0, 76⸱0] | 41⸱0 [20⸱0, 75⸱0] | 42⸱0 [19⸱0, 76⸱0] |
| **Sex** | | | |
| Female (%) | 219 (57⸱9%) | 288 (61⸱8%) | 507 (60⸱1%) |
| Male (%) | 158 (41⸱8%) | 178 (38⸱2%) | 336 (39⸱8%) |
| Not answered (%) | 1 (0⸱3%) | 0 | 1 (0⸱1%) |
| **Weight (kg)** | | | |
| Mean (SD) | 82.2 (19⸱8) | 80⸱8 (21⸱9) | 81⸱5 (21⸱0) |
| Median [Min, Max] | 79.4 [46⸱7, 150] | 79⸱4 [24⸱9, 172] | 79⸱4 [24⸱9, 172] |
| Missing (%) | 22 (5⸱8%) | 37 (7⸱9%) | 59 (7⸱0%) |
| **Height (cm)** | | | |
| Mean (SD) | 168 (18⸱2) | 168 (12⸱8) | 168 (15⸱4) |
| Median [Min, Max] | 168 [0, 198] | 168 [0, 201] | 168 [0, 201] |
| Missing (%) | 2 (0⸱5%) | 3 (0⸱6%) | 5 (0⸱6%) |
| **Body mass index (kg/m^2^)** | | | |
| Mean (SD) | 28⸱5 (6⸱19) | 28⸱1 (6⸱55) | 28⸱3 (6⸱39) |
| Median [Min, Max] | 27⸱3 [16⸱5, 53⸱3] | 26⸱8 [9⸱44, 53⸱9] | 27⸱1 [9⸱44, 53⸱9] |
| Missing (%) | 23 (6⸱1%) | 38 (8⸱2%) | 61 (7⸱2%) |
| **COVID infection before study entry** | | | |
| No | 290 (76⸱7%) | 342 (73⸱4%) | 632 (74⸱9%) |
| Yes | 88 (23⸱3%) | 124 (26⸱6%) | 212 (25⸱1%) |
| **Race/ethnicity (%)** | | | |
| American Indian or Alaskan Native | 4 (1⸱1%) | 3 (0⸱6%) | 7 (0⸱8%) |
| Asian | 41 (10⸱8%) | 62 (13⸱3%) | 103 (12⸱2%) |
| Black or African American | 40 (10⸱6%) | 52 (11⸱2%) | 92 (10⸱9%) |
| Hispanic or Latino | 33 (8⸱7%) | 46 (9⸱9%) | 79 (9⸱4%) |
| Missing | 2 (0⸱5%) | 0 | 2 (0⸱2%) |
| Native Hawaiian or Other Pacific Islander | 1 (0⸱3%) | 1 (0⸱2%) | 2 (0⸱2%) |
| Other | 49 (13⸱0%) | 77 (16⸱5%) | 126 (14⸱9%) |
| White | 208 (55⸱0%) | 223 (47⸱9%) | 431 (51⸱1%) |
| Race/ethnicity not listed here | 0 | 2 (0⸱4%) | 2 (0⸱2%) |

MMM, Moderna-only vaccine/booster series; PPP, Pfizer-only vaccine/booster series; SD, standard deviation.
